# Test-retest reliability of a 2-dimensional and 3-dimensional visual assessment of body image disturbance in anorexia nervosa

**DOI:** 10.1101/2023.06.14.23291397

**Authors:** Christina Ralph-Nearman, Armen C. Arevian, Andrew Karem, Tomas F. Llano-Rios, Megan Sinik, Scott Moseman, Jamie D. Feusner, Sahib S. Khalsa

## Abstract

Body image disturbance (BID) is a diagnostic feature of anorexia nervosa (AN), with few reliable visual perceptual or attitudinal markers. *Somatomap* is a 2-dimensional (2D) and 3-dimensional (3D) digital assessment of BID, which has demonstrated utility. Test-retest reliability of *Somatomap* 2D and 3D digital assessment of BID in AN was examined. Fifty-nine inpatient participants with AN performed test-retest by a) outlining body concern areas on a 2D avatar for each independent area of concern; and b) sculpting 23 independent body parts on a randomized 3D avatar to reflect their perceived body size in length and girth. Participants corresponding body parts were physically measured to calculate discrepancy scores (i.e., 3D perceived minus measured values). Regional 2D BID test-retest differences were evaluated using z-scores to generate statistical visual body maps. Test-retest of 3D assessment reliability was evaluated by Intraclass Correlation Coefficient for individual and aggregated body parts. Somatomap 2D demonstrated excellent test-retest reliability with no statistical differences between test and retest z-scores. All 23 body parts on Somatomap 3D demonstrated statistically significant fair-to-excellent test-retest reliability in AN. Regions that are commonly of concern in AN, and combined measures, showed the highest reliability. Results suggest that Somatomap 2D and 3D may provide a reliable perceptual marker of visual BID in AN.

## Introduction

Anorexia nervosa (AN) takes one life every 52 minutes in the United States alone (Deloitte Access Economics, 2020), and disturbance in how an individual perceives and experiences their body size, or body image disturbance, is a key diagnostic characteristic in detecting increased relapse risk of this serious and potentially fatal mental health illness (American Psychiatric Association [APA], 2013; Gardner&Brown, 2014; Keel et al., 2005). However, there are few validated, visual assessments of body image disturbance, which leaves clinicians with few methods to capture more objectively the multi-faceted perceptual, attitudinal, and interoceptive measures of body image disturbance. Reliable tools with low propensity for bias to assess body image disturbance could help elucidate the cognitive-affective, perceptual, and interoceptive mechanisms underlying body image disturbance and may lead to more efficacious eating disorder treatments and reduce relapse.

Current methods to clinically measure body image disturbance most commonly utilize subjective linguistic-based questionnaires (Cash et al., 2002), or visually based silhouettes (e.g., Stunkard et al., 1983; Swami et al., 2008), and avatars (e.g., Letosa-Porta et al., 2005; Tovée et al., 2003) and are typically whole-body measurement approaches. While useful in some settings, these methods often do not allow for multi-level, more objective, or precise measurement of specific and independent body-part flexibility aspects of body image disturbance, including examining specific body part perceptual distortions and discrepancies (e.g., perceiving ones’ abdomen larger than its objective size) and specific body part-focused concerns (e.g., having body concerns with ones’ thighs) (e.g.,Gardner&Brown, 2014). To address these gaps, the digital tool *Somatomap* was developed (Ralph-Nearman et al., 2019; Ralph-Nearman et al., 2021).

To date, specific patterns of body image concern and body part size estimation accuracy has been identified and mapped using Somatomap on 2-dimensional (2D) and 3-dimensional (3-D) avatars for fashion models as compared to those for nonmodels (see Ralph-Nearman et al., 2019). Results demonstrated that professional fashion models, who have consistent exposure to their body part size through their employment and fitting clothing, were significantly more accurate in predictions than nonmodels in estimating their current perceived body part size compared to the actual measurements of the corresponding body part. Interestingly, the models were less accurate in sculpting their thigh girth on 3D compared with their actual thigh measurement (body discrepancy score), which was also their most frequently endorsed body area of the concern on 2D. This suggested potential for the application of Somatomap in assessment of perceptual body image distortion in eating disorders. In a recent study using this tool, body image disturbance in inpatient individuals with AN were examined (Ralph-Nearman et al., 2021). Results suggested that Somatomap 2D and 3D could pinpoint and visually map independent body part body image disturbances and perceptual distortions. Specific body image disturbances and part perceptual distortions were shown to be related to illness severity in inpatient individuals diagnosed with AN, as well as exhibiting more perceptual discrepancy and dissatisfaction with the body than healthy comparisons. The next step in the process is to test Somatomap 2D’s and 3D’s potential as a digital assessment tool for reliably measuring body image disturbance in clinical populations, such as in individuals with eating disorders, by conducting test-retest reliability with a clinical population.

## Main Aims

Accordingly, the current study aimed to examine test-retest reliability of measuring body image distortion and visual mapping body concerns with Somatomap in an inpatient clinical sample diagnosed with AN. First, the statistical differences were measured between the test and retest for their selected areas of body concern on the 2D avatar. Second, the intraclass Correlation Coefficient (ICC) were measured between the test and retest for the body discrepancy (current perceived minus actual measurements) for each of the 23 separate body parts, as well as the aggregated girth, length, and all body parts on the 3D avatar. We hypothesized that Somatomap 2D would demonstrate test-retest reliability, as demonstrated by the absence of statistically significant visual 2D mapping differences between test and retest. We also predicted that Somatomap 3D would be statistically reliable for body parts and aggregated body discrepancies.

## Method

### Participants

Approval was obtained from the appropriate Institutional Review Board, and procedures used in this study adhere to the tenets of the Declaration of Helsinki. After approval from the Western Institutional Review Board, 59 female inpatients with primary diagnosis of AN with ages between 13 and 38 (*M*=21.08, *SD*=6.04) were recruited from an inpatient eating disorder program. Fifty participants identified as White/Caucasian, and nine participants identified as American Indian/Alaska Native, Hispanic/Latino descent, or ‘something else.’

### Test-retest of Body Image Disturbance via Somatomap 2D and 3D

Each participant provided written informed consent, and for any participants under the age of 18 years, legal guardians also provided consent. Test-retest reliability was assessed on two occasions within the same visit, with ∼30 minutes between the 2D and 3D test and retest to avoid confounds due to inpatient intensive treatment. The order that 2D or 3D was presented to participants on the Chorus digital platform (Arevian et al., 2020; i.e., 2D first and then 3D second, or 3D first and then 2D second) was counterbalanced between participants, and the retest followed the same order as the test for each participant.

For Somatomap 2D, participants first indicated a single area of body concern by circling the area on a digital 2D human avatar. Participants then selected specific types of concerns and emotions related to that independent area of concern. If an individual had more than one area of concern, then the procedure was repeated for each individual area of body concern (see Ralph-Nearman et al., 2021 for more details). Test-retest measured the statistical difference between areas of concern in 2D between tests using z-scores (see Figure 1.a. in which hot and warm heatmap colors would demonstrate any statistically significant concerns differences between test and retest results).

**Figure 1.a.**
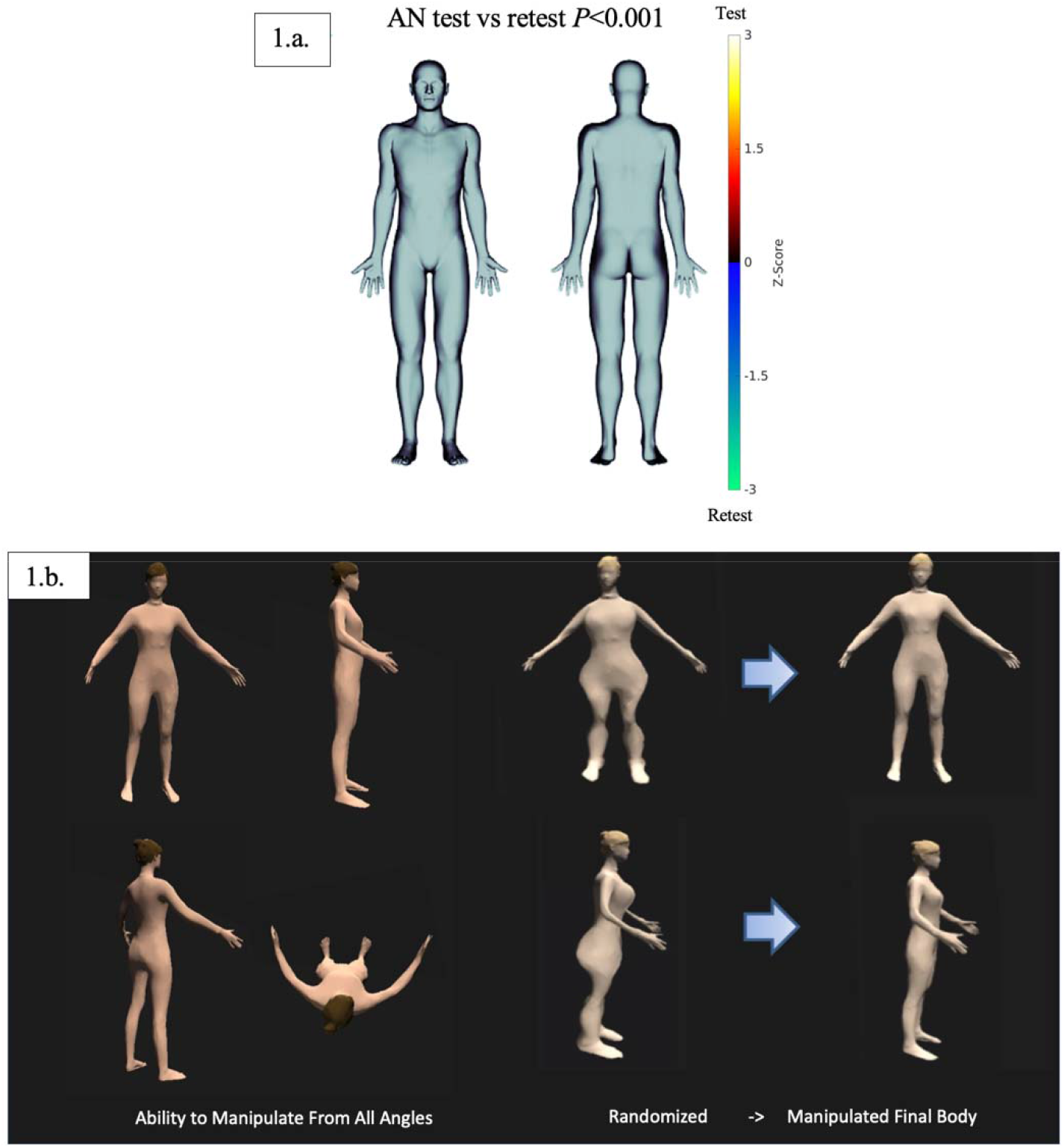
Somatomap 2D: Statistical body map evaluating differences in body image concerns between AN’s test and retest. Any statistically more body image concerns between the retest and test results would have appeared in cool or warm colors on the body maps; *P* < 0.001 statistical threshold. Thus, *lack* of any cool or warm colors indicates the lack of statistically significant differences between test and retest results; **1.b**. Somatomap 3D: Participants use sliders to sculpt the 23 girth/length body parts with sliders from a randomized body to their current perceived body size and shape from all angles.

For Somatomap 3D, participants first personalized the avatar in terms of hair and skin color using sliders. Participants then adjusted the length and girth of 23 body parts, starting from each of the originally presented and randomized sizes and shapes, to represent their perceived current body most accurately (see Ralph-Nearman et al., 2021 for more details). They were able to adjust the body parts from different angles, such as looking down on their body or from the side or back (see Figure 1.b.). Then corresponding physical measurements were obtained for each participant. The discrepancy between participants perceived current and actual measurements was calculated (perceived minus actual measurements). Test-retest assessment relied on the Intraclass Correlation Coefficient (ICC) between the test and retest of 3D for discrepancies for girths and lengths of the separate body parts (i.e., neck girth, neck length, shoulders width, bust girth, chest girth, biceps girth, upper arms length, forearms girth, lower arms length, wrists girth, hands girth, hands length, torso length, waist, stomach form, hips, thighs girth, thighs length, calves girth, calves length, ankles, feet width, and feet length), and aggregated girths, lengths, and all body parts together (see Figure 1.b.).

### Statistical Analysis

Sample size in the current study was based upon prior papers, such as Ralph-Nearman et al., 2021, with an AN sample (*n*=55) with medium to large effect sizes (Cohen’s *F*: 0.25 – 0.5) and Arifin (2022) sample size calculator for ICCs was used to calculated if sample size of *N*=59 would be sufficient. With the current sample size at a minimum reliability of 0.5, expected reliability of 0.8, we estimated over 90% power to detect differences (i.e., extremely high power to detect ICC’s in individual body areas) at α=.01. With 2D our previous studies have shown sensitivity in detecting body perception differences with group sizes as low as *n*=15, therefore we were confident that a sample with greater than three times the sample size will be adequate to detect statistical body map differences.

For Somatomap 2D, Matlab (Mathworks, Inc.) was used to evaluate the differences between the test and retest body concern maps at the statistical level of our previous Somatomap 2D results with individuals diagnosed with AN (*P*<.001; see Figure 1.a.) by calculating z-scores and performing permutation tests for each pixel with a 6-pixel cluster correction and smoothing, as described in prior Somatomap procedures (e.g., Ralph-Nearman et al., 2021).

For Somatomap 3D, the current body discrepancy scores for each of the 23 parts (see Figure 1.b.) were calculated by subtracting the actual measured body part from its corresponding manipulated current perceived body part for the test and the retest. Then the test-retest reliability was assessed by ICC with 95% confidence intervals (CI; absolute-agreement 2-way mixed-effect model; see Koo&Li, 2016) for separate body parts and aggregated girths, lengths, and all body parts.

## Results

### Somatomap 2D test-retest results

Statistical body map analysis results for Somatomap 2D showed no statistical differences between AN participants’ perception of their body concerns on the 2D visual body map between the test and retest z-scores (*P <* 0.001), as shown by the lack of cool or hot colors on the 2D visual body map (see Figure 1.a.).

### Somatomap 3D test-retest results

Test-retest ICCs for Somatomap 3D (see Figure 1.b.) were significant for individual as well as aggregated body parts for current body discrepancy (current perceived minus actual measurements) (see Table 1). Common AN girth-related concerns (e.g., neck, bust, waist, hip, thigh) had the highest reliability (see ICCs≥.75 in bold in Table 1), and aggregate girth body discrepancy had the highest reliability overall (ICC=.93).

**Table 1.** Test-retest ICCs for 23 Body Parts Somatomap 3D Current Body Discrepancy

| Body Part Discrepancy (Girths) | ICC (95% CI)** | p-value* | Reliability |
| --- | --- | --- | --- |
| Neck Girth | **.88 (.79-.93)** | <.001 | Good |
| Shoulder Width | **.84 (.74-.91)** | <.001 | Good |
| Bust Girth | **.86 (.76 -.92)** | <.001 | Good |
| Chest Girth | **.81 (.68-.89)** | <.001 | Good |
| Bicep Girth | **.90 (.83-.94)** | <.001 | Excellent |
| Forearm Girth | **.82 (.69-.90)** | <.001 | Good |
| Wrist Girth | .55 (.24-.73) | .002 | Moderate |
| Hand Width | .70 (.49-.82) | <.001 | Moderate |
| Waist Girth | **.83 (.71-.90)** | <.001 | Good |
| Stomach Form | **.79 (.64-.87)** | <.001 | Good |
| Hip Girth | **.79 (.65-.88)** | <.001 | Good |
| Thigh Girth | **.85 (.72-.92)** | <.001 | Good |
| Calf Girth | .74 (.57-.85) | <.001 | Moderate |
| Ankle Girth | .43 (.04-.66) | .018 | Fair |
| Feet Width | .69 (.48-.82) | <.001 | Moderate |
| **Body Part Discrepancy (Lengths)** |  |  |  |
| Neck Length | **.77 (.61-.86)** | <.001 | Good |
| Upper Arm Length | .73 (.54-.84) | <.001 | Moderate |
| Lower Arm Length | .68 (.46-.81) | <.001 | Moderate |
| Hand Length | **.77 (.62-.86)** | <.001 | Good |
| Torso Length | .70 (.49-.82) | <.001 | Moderate |
| Upper Leg Length | **.75 (.57-.85)** | <.001 | Good |
| Lower Leg Length | .60 (.33-.76) | <.001 | Moderate |
| Feet Length | .69 (.48-.82) | <.001 | Moderate |
| **Body Discrepancy (Aggregated)** |  |  |  |
| Girths | **.93 (.86-.96)** | <.001 | Excellent |
| Lengths | .74 (.56-.85) | <.001 | Moderate |
| **All Body Parts** | **.89 (.81-.93)** | <.001 | Good |

Note. \*Indicates all test and retest were significantly related; \*\*Bold denotes ICCs $\geq .75$ .

## Discussion

The current study examined the test-retest reliability of an innovative digital 2D and 3D avatar-based assessment for visually mapping multi-dimensions of body image disturbance in individuals diagnosed with AN in an inpatient eating disorder treatment center using. As hypothesized, Somatomap 2D demonstrated strong reliability, displaying no statistical body concern visual mapping differences between test and retest at *P* < 0.001 statistical threshold (see Figure 1.a.). Also, Somatomap 3D results showed that all test and retest tests were significantly related. Further, there were good to excellent ICCs on girth-related body parts, which are common concerns for individuals with AN, such as the neck, bust, waist, hip, and thigh (see Table 1). Importantly, unlike many body image disturbance assessments which do not allow for individual body part assessment and/or remove the neck and head from the measure (e.g., Swami et al., 2008), the Somatomap tool is uniquely able to independently and reliably assess these independent body areas that are of common concern for individuals with AN. Interesting to note is that girth-related measures on Somatomap 3D showed higher ICCs than length-related measures. Whether this is a general phenomenon in humans, or specific to AN, will need to be determined in future studies.

Prior studies using Somatomap have demonstrated that this digital assessment tool is able to detect body concern and body size estimation accuracy differences between fashion models and non-models (Ralph-Nearman et al., 2020), as well as specific body concerns and degree of perceptual distortion being significantly related to illness severity in AN (Ralph-Nearman et al., 2021). The current results reinforce and build upon this previous work, demonstrating that Somatomap 2D and 3D may provide a reliable marker of visual body image disturbance in AN and may have utility in longitudinal clinical and research assessments.

### Limitations, Strengths and Future Directions

This study had some limitations, which inform future studies. The current test-retest was performed with females diagnosed with AN who were in an inpatient facility, so future studies may examine more diverse clinical samples, in addition to general population, and non-treatment seeking samples. Further, Somatomap currently allows the personalization of the 3D avatar, including the selection of hair and skin color, so this tool is also well suited to test more ethnically and racially diverse samples than were included in this study for generalizability. Another limitation is that we do not yet know the rate at which body image concerns, disturbances and perceptual discrepancies change in healthy or AN populations. To avoid any confounds due to intensive therapeutic treatments within the inpatient ED setting, the current study utilized participants who were newly admitted into an inpatient ED treatment center and were tested and retested on two occasions within the same visit. However, it is theoretically possible that ∼30 minutes between the test and retest for these measures may not be the best timeframe to assess the reliability of the tool, if one’s body perception or body image concerns change very quickly. Future studies may examine a wider range of moment-to-moment changes in body experience disturbances.

### Conclusion

The present study demonstrates the statistically significant test-retest reliability of Somatomap 2D and 3D for comprehensively assessing body image disturbance in AN. The visual mapping methods employed through this digital multi-platform assessment have demonstrated the ability to reliably capture body image disturbance for individual body parts in AN. There is a growing need for effective, more objective, and remote digital assessments and treatments for eating disorders. The ability for Somatomap 2D and 3D to reliably and effectively measure and visually map independent body concerns, perceptual distortions, and body disturbance, which we have previously shown to be associated with illness severity in AN, may have important and useful applications for both research and clinical settings.

## Data Availability

All data produced in the present study may be obtained by writing first author with reasonable request.

## Declarations

### Ethical Approval

This study was approved by the Western Institutional Review Board, and procedures used in this study adhere to the tenets of the Declaration of Helsinki. All methods and procedures were carried out in accordance with relevant guidelines and regulations. Prior to participating, participants provided written informed consent (Trial Registration: ClinicalTrials.gov #NCT03758326) and data collection procedure is detailed in manuscript.

### Authors’ contributions

C.R.N. analyzed data, wrote the main manuscript text, prepared all figures and tables. M.S. organized data, T.L.R. and A.K. provided cluster for analyses. All authors read manuscript and provided feedback/edits.

### Availability of data and materials

Data may be obtained by writing first author with reasonable request.

## Acknowledgments

We would like to thank Rachel Lapidus. M.S., Danielle Deville, M.S., Olivia Shadid, M.D., Abigail Kimball, B.S., and Alexandra Weindel, B.S., for assistance with data collection, Beth Persac, L.M.F.T. and all the clinicians, therapists, and dieticians at the Laureate Eating Disorders Program for participant coordination, Catherine Wilkerson, Joseph Mango, and Rajay Kumar for technical support, Shane Nearman for graphic creation, and our human models who provided body scans to generate the 3D avatars.

